# Long COVID Citizen Scientists – Developing a needs-based research agenda by persons affected by Long COVID

**DOI:** 10.1101/2021.12.08.21267181

**Authors:** Sarah Ziegler, Alessia Raineri, Vasileios Nittas, Natalie Rangelov, Fabian Vollrath, Chantal Britt, Milo A Puhan

## Abstract

**Objectives:** To identify research priorities of people with Long COVID.

**Design:** Citizen science study following an iterative process of patient needs identification, evaluation and prioritization. A Long COVID Citizen Science Board and a Long COVID Working Group were formed.

**Setting:** Online participation with four activities: three remote meetings and one online survey. First, board members identified needs and research questions. Second, working group members and persons affected by Long COVID evaluated the research questions on a 1-5 Likert scale using an online survey. Then the board gave feedback on this evaluation. Finally, board members set the priorities for research through voting and discussion.

**Participants:** 28 Long COVID Citizen Science Board members: 21 with Long COVID, and 7 with myalgic encephalomyelitis/chronic fatigue syndrome (ME/CFS). 30 Long COVID Working Group members: 25 with Long COVID, 4 ME/CFS patients, and 1 relative. 241 online survey respondents: 85.5% with Long COVID, 14.5% ME/CFS patients, and 7.1% relatives.

**Main outcome measures:** Prioritization of Long COVID-related research questions.

**Results:** 68 research questions were generated by the board and categorized into four research domains (medicine, health care services, socio-economics, and burden of disease) and 14 subcategories. Their average importance ratings were moderate to high and varied from 3.41 (SD = 1.16) for sex-specific diagnostics to 4.86 (SD = 0.41) for medical questions on treatment. Five topics were prioritised: “treatment, rehabilitation and chronic care management”, “availability of interfaces for treatment continuity”, “availability of healthcare structures”, “awareness and knowledge among professionals”, and “prevalence of Long COVID in children and adolescents”.

**Conclusions:** To our knowledge, this is the first study developing a citizen-driven, explicitly patient-centred research agenda with persons affected by Long COVID, setting it apart from existing multi-stakeholder efforts. The identified priorities could guide future research and funding allocation. Our methodology establishes a framework for citizen-driven research agendas, suitable for transfer to other diseases.

**What is already known on this topic:**

- Long-term health consequences following acute SARS-CoV-2 infection, referred to as post COVID-19 condition by WHO or as Long COVID by affected people, are increasing, with population-based prevalence estimates for adults at around 20%.
- Long COVID is associated with fatigue, shortness of breath, cognitive impairment, sleep disorders, pain and other health problems.
- There is little evidence about the needs of persons affected by Long COVID, and it is not clear which research questions should be prioritized to address these needs and help patients and their clinicians make informed and shared decisions about their care.

**What this study adds:**

- Research priorities most important to persons affected by Long COVID are “treatment, rehabilitation and chronic care management”, “availability of interfaces for treatment continuity”, “availability of healthcare structures”, “awareness and knowledge among professionals”, and “prevalence of Long COVID in children and adolescents”.
- Our study may serve as model for a new framework for patient-centred citizen-driven research agendas and as guidance for funding bodies.

## Introduction

The number of people reporting long-term health consequences following acute SARS-CoV-2 infection is increasing. Population-based prevalence estimates for adults centre around 20% but there is large variability across studies depending on the population studied, the length of the follow-up period, and the chosen definition of Long COVID.^1–3^ People commonly report fatigue, shortness of breath, cognitive dysfunction, sleep disorders, pain or inability to return to work or normal social life.^2^ Such long-term consequences are referred to by the World Health Organization (WHO) as post COVID-19 condition. As such, *“post COVID-19 condition occurs in individuals with a history of probable or confirmed SARS-CoV-2 infection, usually 3 months from the onset of COVID-19 with symptoms that last for at least 2 months and cannot be explained by an alternative diagnosis”*.^4^ While the WHO uses the term “post COVID-19 condition”, those affected have been using the term “Long COVID”, which seems to be broadly accepted and widely used across media platforms and by the general population.^5^ As our research directly involved affected people, we decided to consistently use the term “Long COVID”.

Since Long COVID is a novel syndrome, evidence to guide clinical decision-making remains scarce. Little is known about Long COVID’s pathogenesis and risk factors, the benefits or harms of potential treatment options, as well as about best care models to minimize under- and overtreatment. Consequently, funding bodies worldwide have decided to allocate several billions to research on Long COVID.^6–8^ Calls for research proposals are often generated by medical experts and sometimes policy makers, predominantly targeting biological processes, diagnostic and prognostic indicators, as well as therapies.

To efficiently allocate funding resources, its essential to identify research priorities that not only reflect the questions of the medical and scientific community, but also the needs of the patients. While various multi-stakeholder efforts included persons affected by Long COVID in their research priority setting process,^2,9,10^ it is not clear neither how much weight their voice carried over expert opinions, nor if their needs were actually met. In fact, no systematic identification of patients’ needs has been conducted so far, nor has been clarified which research questions should be prioritized to meet those needs. Therefore, our aim was to fill this gap and define research priorities solely from the perspective of persons affected by Long COVID.

## Methods

Our goal was to systematically assess the needs of persons affected by Long COVID, their families, as well as myalgic encephalomyelitis/chronic fatigue syndrome (ME/CFS) patients. Based on this, and always led by those affected, we aimed to derive the most relevant research priorities. For this purpose, we first composed a research team of scientists (VN, MP, AR, SZ), Long COVID network collaborators (NR, FV), and a patient scientist (CB). The research team managed and coordinated the study process without actively taking part in the research priority setting.

### Citizen Scientists

To identify research priorities most important to persons affected by Long COVID we followed an iterative and participatory citizen-science approach including collaborative and co-created participation (Figure 1).^11^ For this purpose, we built two different types and intensities of participation: A Long COVID Citizen Science Board and a Long COVID Working Group. For both, citizen scientists were recruited online via the Altea Long COVID Network platform (https://www.altea-network.com/en/) that is funded by the Federal Office of Public Health and other supporters and Long Covid Switzerland’s Facebook group. Citizen scientists were eligible if they were i) affected by Long COVID, ii) relatives of a person affected by Long COVID, or ii) ME/CFS patients. We decided to include relatives because they are closely involved and often also strongly affected. We also invited ME/CFS patients, who share several common symptoms with Long COVID affected people, to include a long-term perspective, which may still be missing among the other participants. Registration was open from March 25 to April 18, 2021 and interested parties could sign up for the citizen science board, the working group, or both.

**Fig 1.**
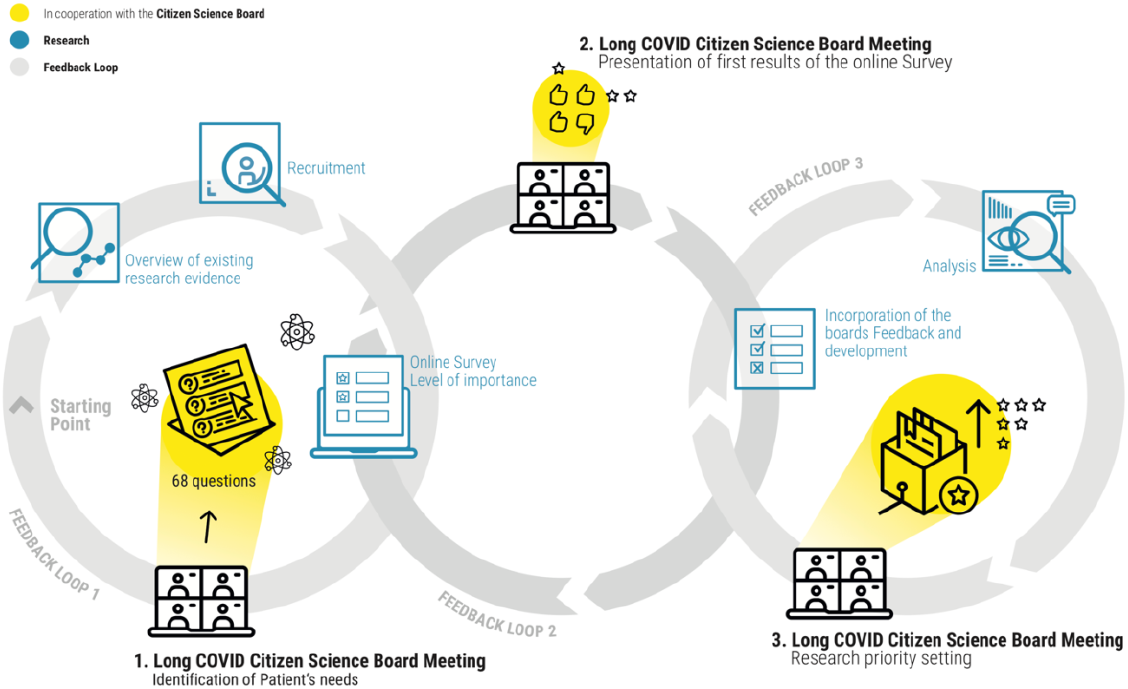
Process and development of a citizen-driven research project to develop research priorities most important to persons affected by Long COVID.

#### Long COVID Citizen Science Board

We aimed to recruit a citizen science board consisting of at least 30 selected citizen scientists: 20 Long COVID patients, five relatives, and five ME/CFS patients. In order to reach a balanced board, two team members (SZ; CB) independently selected eligible citizen scientists based on age, sex, disease severity, and motivation. Consensus was reached through discussion. Remaining interested parties were put on a waiting list or, if desired, assigned to the Long COVID Working Group. Participation in the Long COVID Citizen Science Board was following co-creation principles throughout the research process including three iterative feedback-loops (online meetings), from problem definition to research question formulation and final prioritization (Figure 1).

#### Long COVID Working Group

Participation in the Long COVID Working Group was contributory and collaborative but in a less demanding and non-binding manner. As such citizen scientists in the working group were not involved in the problem definition, nor the research question formulation and did not participate in online meetings. Instead, they were involved in shaping research priorities by participating in the online survey.

### From peoples’ needs to research priorities

For the purpose of our study, we followed a consensus-oriented process using acceleration room techniques, which are goal-oriented digitally supported group processes. We have successfully used this iterative and participatory technique before.^12^ To this end, we spread the entire research priority setting process over four main activities, including three iterative feedback-loops, which included three Citizen Science Board meetings, and one online survey (Figure 1).

#### 1) Long COVID Citizen Science Board Meeting 1: Needs identification

With the first citizen science board meeting (held online on May 5, 2021, via ZOOM) we aimed to 1) get to know each other, 2) provide a lay summary of existing evidence on Long COVID, 3) exchange experiences about Long COVID, and 4) identify patients’ needs. For this purpose, we structured the meeting into plenary sessions and breakout groups using MURAL. These acceleration room techniques allowed participants to table their thoughts personally or anonymously on the MURAL board that was serving as whiteboard, structuring the discussions and automatically generating a written document.

After the first board meeting, we structured the identified participants’ needs into overall research domains. In a second step, we translated all identified and categorized needs into research questions. Consensus was reached through multiple rounds of discussion and subsequent revisions among the research team. Since the aim of these questions was to guide future research and funding allocation, we explicitly derived general and not specific clinical research questions using the PICO framework.^13^

We then sent the list of all formulated research questions to the members of the Long COVID Citizen Science Board, asking for review and revisions.

#### 2) Online Survey: Needs evaluation

To ensure that the identified research questions represented the needs of persons affected by Long COVID and did not simply reflect the needs of the board, we evaluated the questions through an anonymous online survey conducted between June and September 2021. For each research question, participants could rate the level of importance from 1 (not important) to 5 (very important). The link to the online survey was shared with the Long COVID Working Group, Altea Long COVID Network platform and Long Covid Switzerland’s Facebook group.

#### 3) Long COVID Citizen Science Board Meeting 2: Needs evaluation feedback

In a second citizen science board meeting (held online on July 9, 2021, via ZOOM) we aimed to 1) present interim results of the online survey, and 2) conduct a second feedback loop on the research questions. After this second meeting, we made final revisions on the research questions and analysed the results of the completed online survey, stratified by overarching research domains and subcategories.

#### 4) Long COVID Citizen Science Board Meeting 3: Needs prioritization

In the third citizen science board meeting (held online on October 1, 2021, via ZOOM) we aimed to 1) present and discuss the final survey results, and 2) vote on research priorities. Prior to the meeting, each participant was provided with the survey results, the list of research questions and their categorization into overarching research domains and subcategories. Before discussing the survey results, we asked each participant to select the three research subcategories most important to them. In a second step, we compared the results of the boards’ voting with the survey results. Consensus on the final selection of the research priorities was reached within the board through discussion. Finally, we reprocessed the results of the third meeting and forwarded the identified research priorities to all members of the Long COVID Citizen Science Board, as well as to the Long COVID Working Group for final feedback.

## Results

In total 66 people signed up to contribute as citizen scientists: Two for the board only, 18 for the working group only and 46 for both. From the 46 people who signed up for both groups 33 were selected for the board, 12 were selected for the working group, and one person got rejected due to missing information about the motivation. Of the two people interested in the board only, one was on the waiting list and the other person got rejected due to missing information about the motivation. All 18 people interested in the working group only were selected for the working group.

### Long COVID Citizen Science Board

In total 33 people were selected for the board. Of these, two did not provide any contact details and three dropped out. Finally, the Long COVID Citizen Science Board consisted of 28 citizen scientists (median age 50, age range 32-78 years), 22 were female. 21 board members were affected by Long COVID and seven were ME/CFS patients.

### Long COVID Working Group

In total 30 people were selected for the working group. Of these, two thirds (median age 47, age range 29-78 years) were female. 24 working group members were affected by Long COVID, one member was a relative, and five were ME/CFS patients.

### Identification, evaluation and prioritization of the needs

#### 1) Citizen Science Board Meeting 1: Needs identification

The Long COVID Citizen Science Board identified numerous needs related to Long COVID research. From these, we formulated 68 research questions considering the broader scientific context. The questions were allocated to four overarching research domains and 14 subcategories. Some contributions from the first board meeting could not be formulated as research questions, as they were mostly requests, questions, and concerns that required already established regulatory and infrastructural processes (e.g. legal or clinical guidelines). These were listed in a separate category defined as “further questions” (Supplement 1). An overview of the four emerged research domains and 14 subcategories is presented in Figure 2.

**Fig 2.**
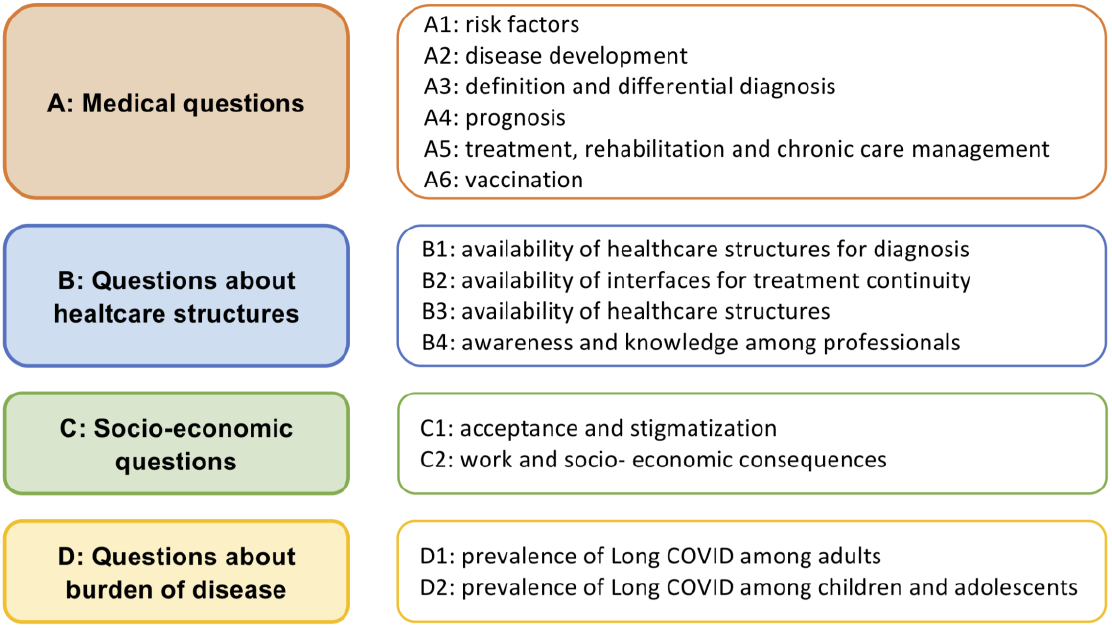
Overview of the research themes and subcategories identified by the Long COVID Citizen Science Board.

The first domain, named “medical questions”, consisted of 32 research questions grouped into six subcategories (A1–A6): A1 “risk factors”, A2 “disease development”, A3 “definition and differential diagnosis”, A4 “prognosis”, A5 “treatment, rehabilitation and chronic care management”, and A6 “vaccination”. The second domain, “questions about healthcare structures”, included 20 research questions grouped into four subcategories (B1–B4): B1 “availability of healthcare structures for diagnosis”, B2 “availability of interfaces for treatment continuity”, B3 “availability of healthcare structures”, and B4 “awareness and knowledge among professionals”. The third domain named “socio-economic questions” included eight research questions split into two subcategories (C1–C2): C1 “acceptance and stigmatization”, and C2 “work and socio-economic consequences”. Finally, the fourth domain, “burden of disease” included eight research questions split in two subcategories (D1–D2): D1 “Prevalence of Long COVID among adults”, and D2 “Prevalence of Long COVID among children and adolescents”.

#### 2) Online Survey: Needs evaluation

In total, 241 people (83.8% women, mean age 46 years) completed the online questionnaire on the level of importance of the 68 research questions. Most of the participants were affected by Long COVID (85.5%), 14.5 % suffered from ME/CFS and 7.1% were relatives. For details see Table 1.

**Table 1.** Baseline characteristics of 241 people participating in the online survey on the Level of Importance of 68 research questions about Long COVID.

| **Characteristics and category** | **%(No)** |
| --- | --- |
| Age [Mean, Standard Deviation] | 46.2<br>(12.3) |
| Sex |  |
| Women | 83.8 (202) |
| Men | 16.2 (39) |
| Persons affected by Long COVID | 85.5 (206) |
| Relatives of Long COVID patients | 7.1 (17) |
| ME/CFS patients | 14.5 (35) |

ME=myalgic encephalomyelitis; CFS=chronic fatigue syndrome. Baseline characteristics extracted from online survey conducted between June and September 2021. The link to the online survey was shared with the Long COVID Working Group, Altea Long COVID Network platform and Long Covid Switzerland's Facebook group.

Table 2 shows, in descending order, the participants’ median and average ratings (and standard deviations) of the level of importance of each research question ranging from 1 (not important) to 5 (very important) by domain. In general, none of the 68 research questions were rated as unimportant nor little important and all ratings exceed 3 (neither important nor unimportant). As shown in Figure 3a, there was large variation in the level of importance between the 68 research questions and within the four research domains. While research question A5.1 *“What existing and new therapeutic approaches/treatment methods, depending on diagnosis and severity, are effective to treat Long COVID patients?”* scored highest 4.86 (SD= 0.41), research question A3.3 *“Are there differences in diagnostic criteria between men and women?”* scored lowest 3.41 (SD= 1.16). Despite the large variation within the four research domains, some of the 14 subcategories were perceived to be more important than others. As such, participants consistently rated the subcategory D2 “prevalence of Long COVID in children and adolescents”, A5 “treatment, rehabilitation and chronic care management”, A2 “disease development”, B4 “awareness and knowledge among professionals”, and B3 “availability of healthcare structures” as important or very important (Figure 3b). In contrast, participants indicated with larger variation a lower level of importance to the subcategory A3 “definition and differential diagnosis” and A1 “risk factors”.

**Table 2.**
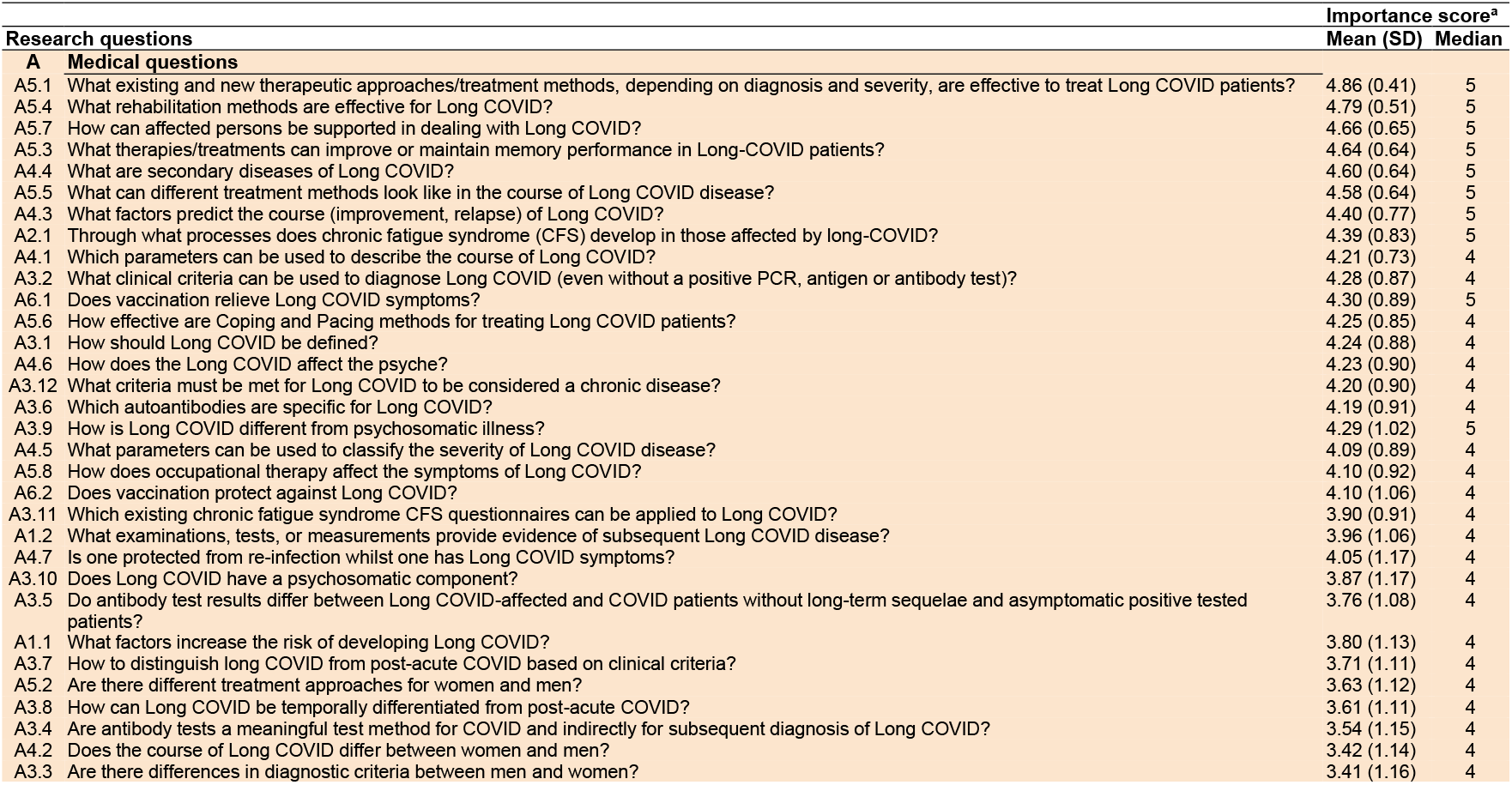
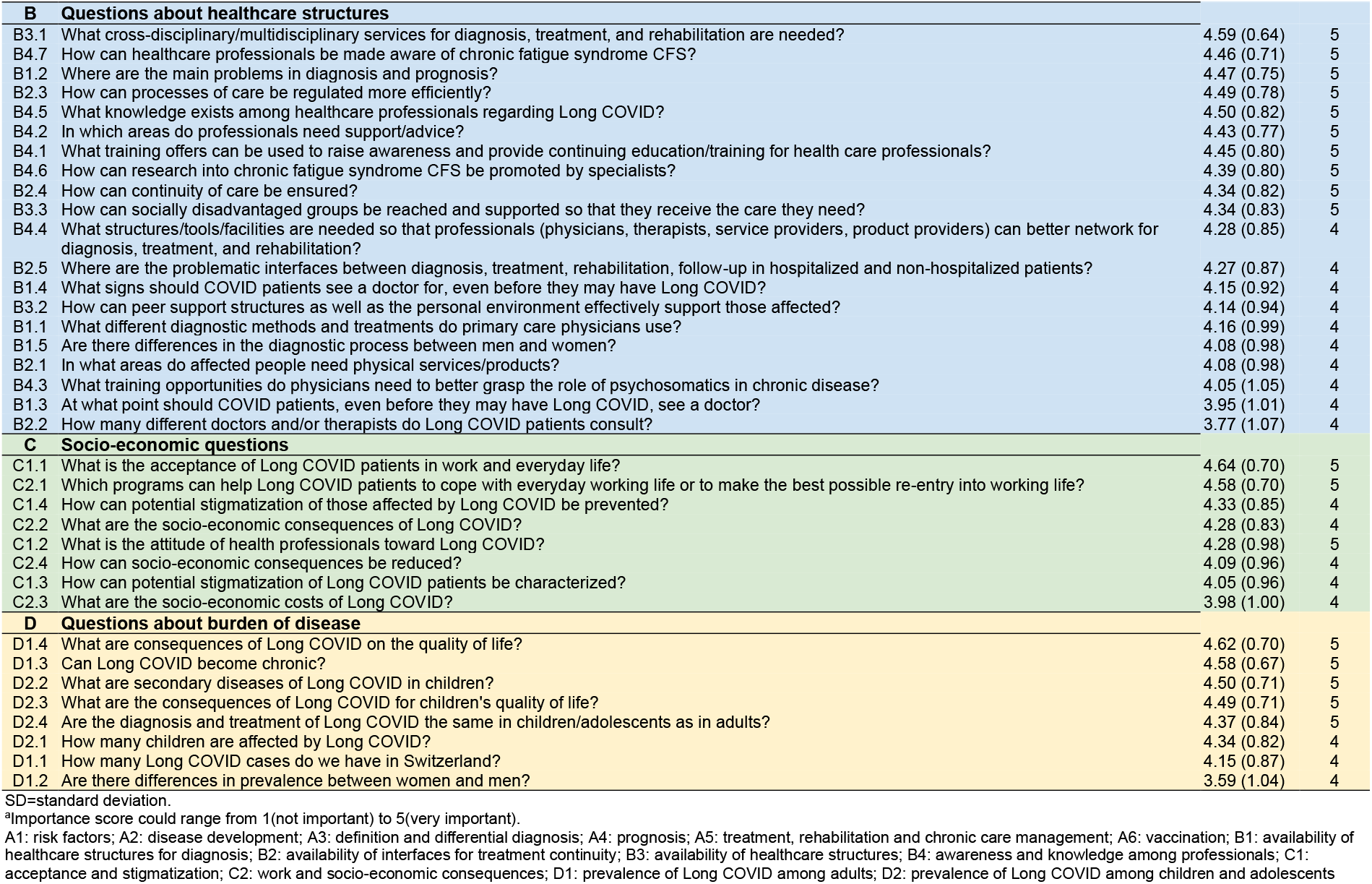
Results from an online survey (June–September 2021) on the level of importance of 68 research questions about Long COVID.

**Fig | 3a.**
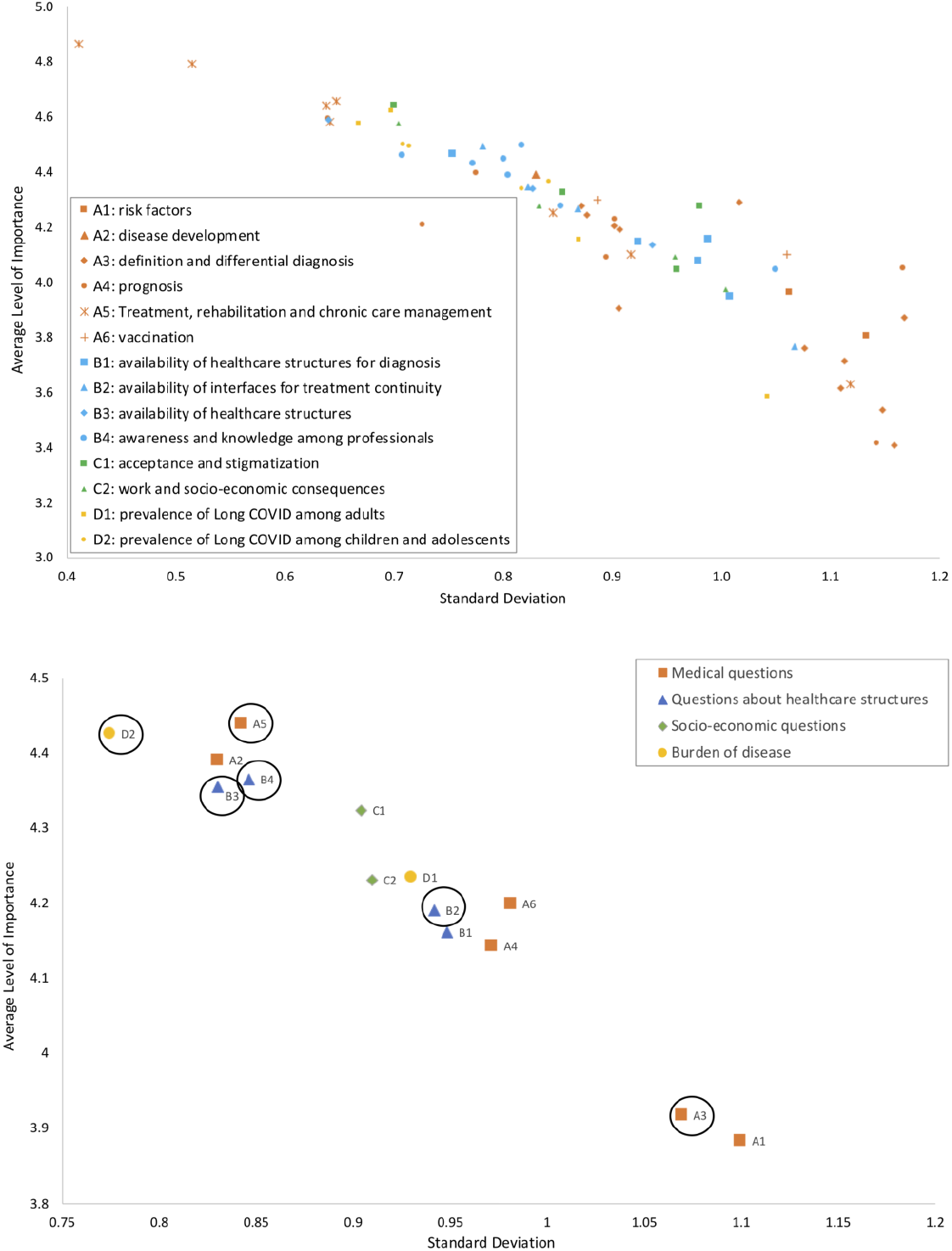
Results of the online survey with 241 participants (June–September 2021) on the level of importance of the 68 research questions identified by the Long COVID Citizen Science Board. Individual research questions are plotted by their average level of importance, ranging from 1 (not important) to 5 (very important) (y-axis) and standard deviation (x-axis). **Fig | 3b**. Results of the online voting on research priorities of the Long COVID Citizen Science Board. The circles refer to the research priorities of the Long COVID Citizen Science Board identified during the third meeting (October 2021).

#### 3) Citizen Science Board Meeting 2: Needs evaluation feedback

The citizen scientists of the board were consistent with the preliminary results of the online evaluation of the 68 research questions. As such, the board agreed that all research domains were generally important with the subcategories D2 “prevalence of Long COVID in children & adolescents”, A5 “treatment, rehabilitation & chronic care management”, A2 “disease development”, B4 “awareness and knowledge among professionals”, and B3 “availability of healthcare structures” being highlighted as important or very important. No further changes were made to the research questions, the four domains or the 14 subcategories.

#### 4) Citizen Science Board Meeting 3: Needs prioritization

Figure 3b shows the research priorities of Long COVID Citizen Science Board members (highlighted in circles), as identified by anonymous online voting during the third meeting. The five identified research priorities were (in random order): A5 “treatment, rehabilitation and chronic care management”, B2 “availability of interfaces for treatment continuity”, B3 “availability of healthcare structures”, B4 “awareness and knowledge among professionals”, and D2 “prevalence of Long COVID in children and adolescents”. The board agreed that subtopic A3 “definition and differential diagnosis” is important and should serve as basis for the remaining subcategories but not as research priority itself.

Most of the board’s research priorities were consistent with the results of the online evaluation. However, the Long COVID Citizen Science Board members put less priority on the subtopic A2 “disease development”, and instead prioritized the subtopic B2 “availability of interfaces for treatment continuity”.

## Discussion

This is the first project to recruit Long COVID citizen scientists in order to identify and prioritise timely, patient-relevant research topics. The five identified research priorities are: A5 “treatment, rehabilitation and chronic care management”, B2 “availability of interfaces for treatment continuity”, B3 “availability of healthcare structures”, B4 “awareness and knowledge among professionals”, and D2 “prevalence of Long COVID in children and adolescents”. Not surprisingly, medical questions such as understanding symptoms and disease progression, underlying causes and treatment emerged as prominent research topics. These topics highly overlap with findings from other research priority setting efforts following multi-stakeholder approaches.^2,9,10^

Being diagnosed in a timely and correct manner seems to be one of the biggest challenges for those affected by Long COVID. The new clinical case definition of the WHO serves as a starting point toward a common understanding of Long COVID and ultimately improved diagnostic procedures.^4,5^ Yet, it remains to be seen to what extent that definition will impact clinical practice. The unspecific and heterogeneous nature of Long COVID symptoms ^1,14^ will likely continue to challenge the development of clear universally accepted diagnostic guidelines and differentiation from other conditions, such as ME/CFS.^15^

Further, in our study, the high average importance of the corresponding research questions indicates that, in addition to a clear diagnosis, persons affected by Long COVID are currently mostly missing adequate treatment options and access to adequate care that meets their multidimensional needs. Indeed, standard medical care is currently insufficient to alleviate the heterogeneous symptom burden of Long COVID. Our results emphasise the importance of appropriate care structures for adequate diagnosis and treatment, including efficiently regulated supply processes, improved continuity of care, and better awareness, as well as knowledge about and understanding of Long COVID among healthcare professionals. This indicates an increased need for integrated multi-disciplinary care structures ^16^ as an integral part of care management, incrementally, over the course of the illness. More research on health services is needed to determine how such approaches can be linked to existing care structures to make efficient use of available resources.

The needs of persons affected by Long COVID are not merely medical, but also include multiple social and psychological elements. Our results show that on an individual level, people were concerned about losing their jobs and potential stigmatization by healthcare professionals, as well as by the broader social environment, including work and family. Evidence on Long COVID-related stigmatization remains scarce, but the lack of adequate healthcare structures may indicate institutional discrimination.^17^ Institutional discrimination occurs when healthcare systems fail to provide the right care to people because of their health condition.^18^ In the case of Long COVID, major drivers are likely the until recently lacking universal definition, as well as the complex diagnostic procedures. For instance, in many countries, people suffering from Long COVID are only entitled to sick leave after confirmed diagnosis.^19^ Further, a recent living systematic review revealed that Long COVID can affect patients’ family life, social functioning, and ability to work.^1^ In line with this, the National Institute for Health and Research found that Long COVID affected patients’ ability to work in 80% of respondents and patients’ family life in 71% of surveyed patients.^2^ The full extent of Long COVID’s socio-economic implications remains to be determined. Despite this, socio-economic questions are not a priority in the research agendas developed by multi-stakeholder approaches.^2,9,10,20^

In our study, the domain “prevalence of Long COVID among children and adolescents” was also deemed important and within this domain, the questions about how many children are affected by Long COVID, whether there are secondary diseases that arise because of Long COVID in children, and how Long COVID affects children’s quality of life. A longitudinal cohort study found that 2-4% of children enrolled in the first and the second wave of the pandemic reported at least one symptom lasting beyond 12 weeks of acute infection.^21^ Long COVID in children and adolescents was prioritized equally strongly only by one multi-stakeholder study known to us.^10^ The Swedish Agency for Health Technology Assessment and Evaluation of Social Services (SBU) also collected research questions on Long COVID in children but did not prioritize them further.^9^ This may be an indication of diverging priorities between those affected by Long COVID and other stakeholders.

### Strengths and Limitations

To our knowledge, this is the first research project that developed a citizen-driven, explicitly patient-centred research agenda, generated by persons affected by Long COVID, in line with current recommendations.^2,20^ This sets our work apart from previous multi-stakeholders’ efforts, in that the research team only managed and coordinated the study process without actively taking part in the research priority setting. The Long COVID Citizen Science Board and the Long COVID Working Group were developed to allow collaborative and co-creative participation, enabling priority setting and research agenda setting solely by participants.

One limitation of our project is that, while invited, relatives did not register to be part of the board, and only few participated in the working group or the online survey. As a consequence, the needs of relatives might be underrepresented. Second, both the recruitment of citizen scientists and of participants for the online evaluation was carried out via the Altea Long COVID Network platform and Long Covid Switzerland’s Facebook group. This means that we have mainly reached people from the German-speaking and French-speaking parts of Switzerland and may have missed people from the Italian-speaking part of Switzerland.

## Conclusions

To the best of our knowledge, this is the first research project that identified Long COVID research priorities using a citizen science approach and solely considering the needs of those affected. The following priorities have been identified and should be included in future studies: “treatment, rehabilitation and chronic care management”, “availability of interfaces for treatment continuity”, “availability of healthcare structures”, “awareness and knowledge among professionals”, and “prevalence of Long COVID in children and adolescents”.

Our methodology can be adapted to other settings and health conditions. It may pave the way towards co-created and patient-centred research agendas, ultimately initiating at least some shift away from the scientific and medical community, to engaging citizens, and holistically acknowledge and embrace their needs.

Ultimately, the five identified research priorities may guide and justify future funding allocation. Indeed, persons affected by Long COVID were at the centre of generating a crucial, timely, evidence base for an emerging, pernicious syndrome that has captured the attention of society and the medical community around the world.

## Public and patient involvement statement

To identify research priorities most important to persons affected by Long COVID we followed an iterative and participatory citizen-science approach including collaborative and co-created participation. For this purpose, we built two different types and intensities of participation: A Long COVID Citizen Science Board and a Long COVID Working Group. In addition, we conducted an online survey to evaluate the needs identified by the citizen scientist. All major decisions about the research priorities have been made by the persons affected by Long COVID.

## Supporting information

Supplementary 1 "Further questions"

## Data Availability

All data produced in the present work are contained in the manuscript.

## Contributors

CB and MAP initiated the project and the preliminary design. AR, CB, MAP, SZ, and VN further developed the design and methods. CB, FV, NR and SZ recruited citizen scientists and online survey participants. AR; CB, FV, MAP, NR, SZ, and VN conducted three citizen science meetings, supervised breakout group discussions and collected data generated during the meetings. SZ managed the communication with the citizen scientists, designed and analysed an online evaluation for the first citizen science board meeting, designed the online survey for evaluating the identified research questions, and collected, managed and analysed the data. SZ wrote the first draft of the manuscript. MAP is the guarantor and accepts full responsibility for the work and the conduct of the study, had access to the data, and controlled the decision to publish. SZ confirms that the manuscript is an honest, accurate, and transparent account of the study being reported; that no important aspects of the study have been omitted; and that any discrepancies from the study as originally planned (and, if relevant, registered) have been explained. We thank the members of the Long COVID Citizen Science Board and the Long COVID Working Group for their valuable contribution to this study.

## Ethics approval

The ethics committee of the canton of Zurich issued a waiver (Req-2021-01415) since the study does not fall under responsibility of the law on human research.

