## Supplementary 1 "Further questions" for "Long COVID Citizen Scientists – Developing a needs-based research agenda by persons affected by Long COVID"

**Supplementary file 1 «Further questions»**

What clinical and laboratory parameters can be used to inform disability insurances and other support?

What tests can be used to measure exercise intolerance in Long COVID patients? Should these be newly developed?

Do GPs know specialized points of contact, or appropriate specialists?

Do GPs know the prevalence/incidence of Long COVID?

What evidence (examination, test results) do Long COVID patients subsequently need?

Are there enough specialized clinics for treatment and rehabilitation?

Are quality criteria defined? Who monitors the quality of the services?

What preconditions have to be met for Long COVID to be recognized as a disease?

Which criteria have to be fulfilled for a health insurance company to cover the treatment cost for Long COVID?

What conditions have to be met for Long COVID patients to receive the best possible coverage via insurance? What do patients have to prove?

Which costs can be covered by insurance? What are the respective requirements?

What conditions have to be met for Long COVID patients to be protected against unemployment?

What preconditions have to be created so that Long COVID becomes a socially recognized syndrome?

What factors can help Long COVID patients to reintegrate into social life?

What testing procedures, equipment, guidelines, and knowledge do GPs need to have in order to diagnose Long COVID?

What are possible empowerment supports for dealing with Long COVID?
